# Predicting Mortality Risk in Patients with COVID-19 Using Artificial Intelligence to Help Medical Decision-Making

**DOI:** 10.1101/2020.03.30.20047308

**Authors:** Mohammad Pourhomayoun, Mahdi Shakibi

## Abstract

In the wake of COVID-19 disease, caused by the SARS-CoV-2 virus, we designed and developed a predictive model based on Artificial Intelligence (AI) and Machine Learning algorithms to determine the health risk and predict the mortality risk of patients with COVID-19. In this study, we used documented data of 117,000 patients world-wide with laboratory-confirmed COVID-19. This study proposes an AI model to help hospitals and medical facilities decide who needs to get attention first, who has higher priority to be hospitalized, triage patients when the system is overwhelmed by overcrowding, and eliminate delays in providing the necessary care. The results demonstrate 93% overall accuracy in predicting the mortality rate. We used several machine learning algorithms including Support Vector Machine (SVM), Artificial Neural Networks, Random Forest, Decision Tree, Logistic Regression, and K-Nearest Neighbor (KNN) to predict the mortality rate in patients with COVID-19. In this study, the most alarming symptoms and features were also identified. Finally, we used a separate dataset of COVID-19 patients to evaluate our developed model accuracy, and used confusion matrix to make an in-depth analysis of our classifiers and calculate the sensitivity and specificity of our model.

## I. Introduction

In late 2019, a novel form of Coronavirus, named SARS-CoV-2 (stands for Severe Acute Respiratory Syndrome Coronavirus 2), started spreading in the province of Hubei in China, and claimed numerous human lives [1]-[3]. In January 2020, the World Health Organization (WHO) declared the novel coronavirus outbreak a Public Health Emergency of International Concern (PHEIC) [4][5]. In February 2020, WHO selected an official name, COVID-19 (stands for Coronavirus Disease 2019), for the infectious disease caused by the novel coronavirus, and later in March 2020 declared a COVID-19 Pandemic [5][6].

Coronavirus is a family of viruses that usually causes respiratory tract disease and infections that can be fatal in some cases such as in SARS, MERS, and COVID-19. Some kinds of coronavirus can affect animals, and sometimes, on rare occasions, coronavirus jumps from animal species into the human population. The novel coronavirus might have jumped from an animal species into the human population, and then begun spreading [7]. A recent study has shown that once the coronavirus outbreak starts, it will take less than four weeks to overwhelm the healthcare system. Once the hospital capacity gets overwhelmed, the death rate jumps [8].

Artificial Intelligence (AI) has been shown to be an effective tool in predicting medical conditions and adverse events, and help caregivers with medical decision-making [9]-[13]. In this study, we proposed a data-driven predictive analytics algorithm based on Artificial Intelligence (AI) and machine learning to determine the health risk and predict the mortality risk of patients with COVID-19. The developed system can help hospitals and medical facilities decide who needs to get attention first, who has higher priority to be hospitalized, triage patients when the system is overwhelmed by overcrowding, and eliminate delays in providing the necessary care. The algorithm predicts the mortality risks based on patients’ physiological conditions, symptoms, and demographic information.

The proposed system includes a set of algorithms for preprocessing the data to extract new features, handling missing values, eliminating redundant and useless data elements, and selecting the most informative features. After preprocessing the data, we use machine learning algorithms to develop a predictive model to classify the data, predict the medical condition, and calculate the probability and risk of mortality.

The rest of this paper is organized as follows: in section II, we will introduce the different methods and model architecture. Discuss each method by providing detailed information about the model, data preprocessing, and challenges that we encountered and the steps to mitigate these challenges, feature selection, and feature extraction. In section III, describe the results and conclusion.

## II. Methods

### A. Dataset

In this paper, we used a dataset of more than 117,000 laboratory-confirmed COVID-19 patients from 76 countries around the world including both male and female patients with an average age of 56.6 [3]. The disease confirmed by detection of virus nucleic acid [3]. The original dataset contained 32 data elements from each patient, including demographic and physiological data. At the data cleaning stage, we removed useless and redundant data elements such as data source, admin id, and admin name. Then, Data imputation techniques were used to handle missing values.

After analyzing the data, we found out that 74% of patients were recovered from COVID-19. To have an accurate and unbiased model, we made sure that our dataset is balanced. A balanced dataset with equal observations for both recovered and deceased patients was created to train and test our model. The data observations (patients) in the training dataset have been selected randomly and they are completely separate from the testing data. Figure 1 shows a high-level architecture of our system.

**Figure 1.**
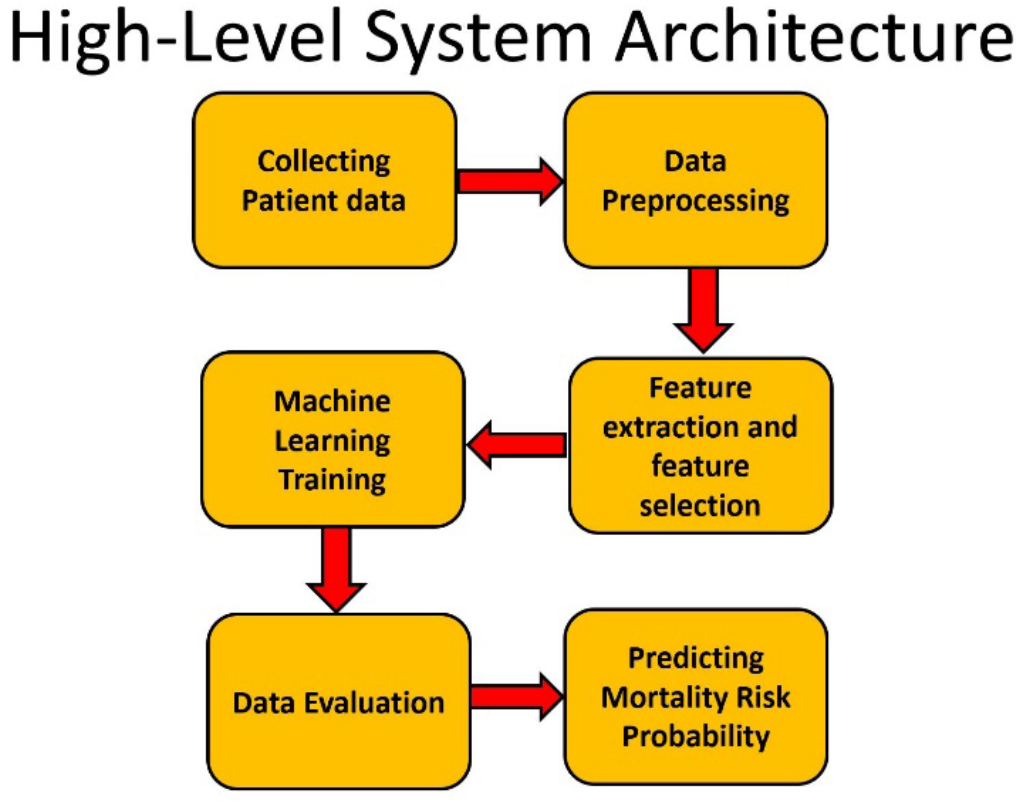
High-level system architecture.

### B. Feature selection

The outcome label contained multiple values for the patient’s health status. We considered patient that discharged from hospital or patients in stable situation with no more symptoms as recovered patients.

A total of 80 features were extracted from *symptoms* and *doctors’ medical notes* about the patient’s health status.

We also extracted additional 32 features from patient’s demographic and physiological data, made it to total 112 features. We consulted with a medical team to make sure that the best features are extracted and selected.

The next step is feature selection. The primary purpose of feature selection is to find the most informative features and eliminate redundant data to reduce the dimensionality and complexity of the model [11]. We used univariate and multivariate filter method and wrapper method to rank the features and select the best feature subset [11]. Figure 2 demonstrates the steps of filter and wrapper method that we used for feature selection.

**Figure 2.**
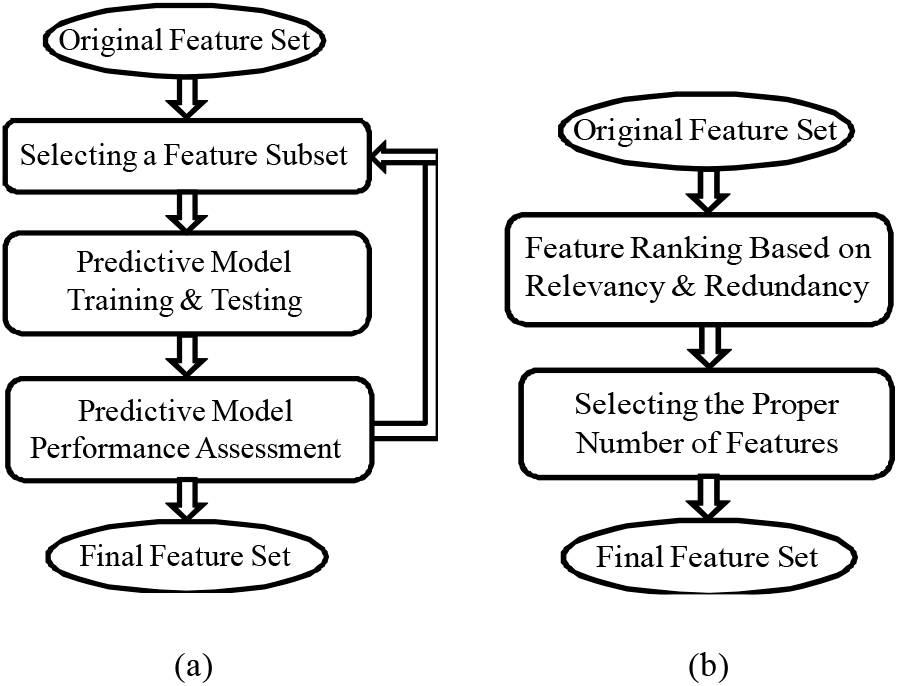
Feature Selection: (a)Wrapper method, (b)Filter method.

Filter methods are very popular (especially for large datasets) since they are usually very fast and much less computationally intensive than wrapper methods. Filter methods use a specific metric to score each individual feature (or a subset of features together). The most popular metrics used in filter methods include correlation coefficient, Fisher score, mutual information, entropy and consistency and chi-square parameters [11].

After applying different filter and wrapper methods, we chose 42 features out of 112 features. Our final feature set includes demographic features such as age, sex, province, country, age, travel history, general medical information such as comorbidities (diabetes, cardiovascular disease, …), and also patient symptoms such as chest pain, chills, colds, conjunctivitis, cough, diarrhea, discomfort, dizziness, dry cough, dyspnea, emesis, expectoration, eye irritation, fatigue, gasp, headache, lesions on chest radiographs, little sputum, malaise, muscle pain, myalgia, obnubilation, pneumonia, myelofibrosis, respiratory symptoms, rhinorrhea, somnolence, sputum, transient fatigue, weakness, etc.

Figure 3 shows the Correlation Heatmap for dataset features. Figure 3-(a) shows the correlation between features and the outcome i.e. mortality risk, and Figure 3-(b) shows the correlation between features. As Figure 3-(a) illustrates, some features like age and chronic diseases (comorbidities) were the top features with high correlation to the patient’s mortality risk.

**Figure 3.**
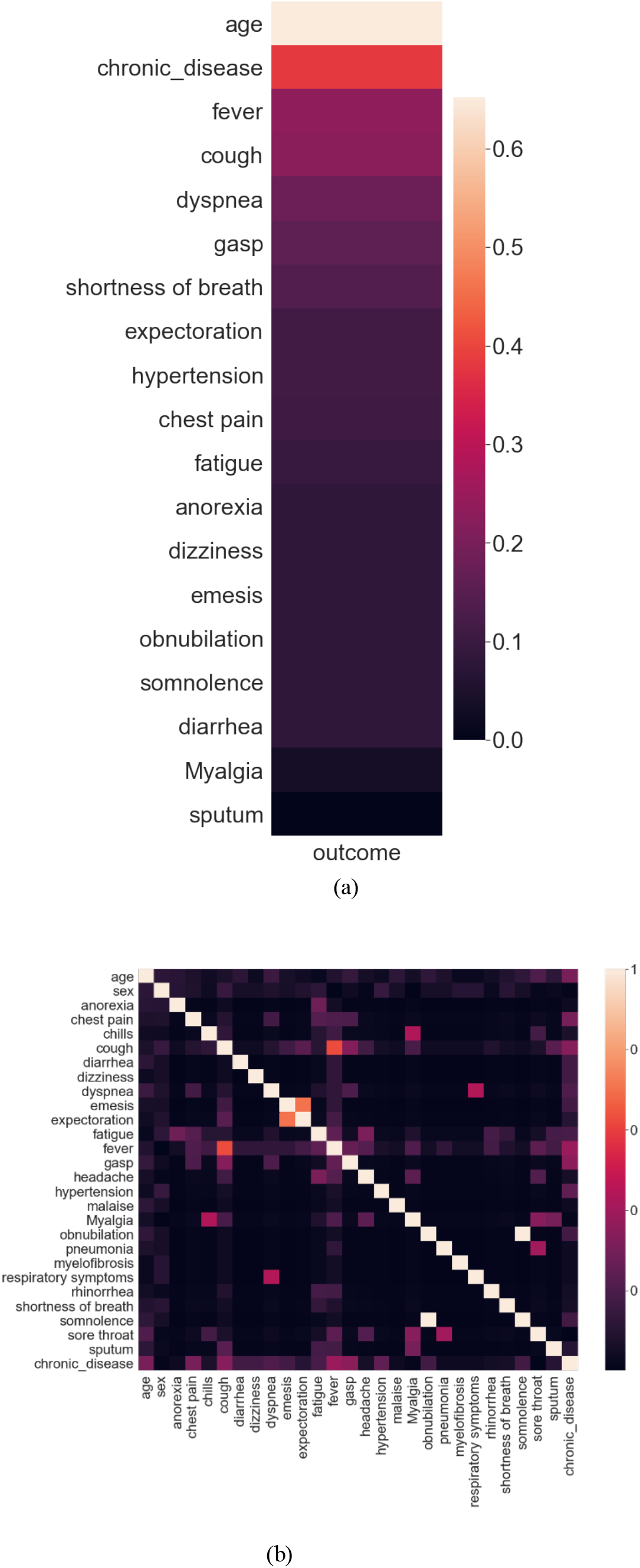
(a) Correlation heatmap for the most correlated features to the mortality risk, (b) Correlation heatmap between features.

### C. Predictive Analytics Algorithms

After selecting the best feature subset, we used various machine learning algorithms to build a predictive model. In this research, we used different algorithms including Support Vector Machine (SVM), Neural Networks, Random Forest, Decision Tree, Logistic Regression, and K-Nearest Neighbor (KNN) [15][16][17].

The Neural Network algorithm achieved the best performance and accuracy. We used grid search to find the best hyper-parameters for the neural network. The best neural network results were achieved with two hidden layers with 10 neurons in the first layer and 3 neurons in the second layer. We used sigmoid function as the hidden layer activation function and used stochastic gradient optimizer, constant learning rate and the regularization rate of alpha = 0.01.

The SVM model was configured with linear kernel, and regularization parameter C=1.0.

The Random Forest algorithm is an ensemble learning method combined of multiple decision tree predictors that are trained based on random data samples and feature subsets [17]. We configured the random forest algorithm with 20 trees in the forest.

### D. Evaluation

We used *10-fold random cross-validation* (with no overlap, with no replacement) to evaluate the developed model. We calculated the Overall Accuracy for all machine learning algorithms to compare. Also, we generated Receiver Operating Characteristic (ROC) curves for every algorithm, and calculated the Area Under Curve (AUC) and Confusion Matrix. Again, we made sure that there is no overlap (no common patient) between training and testing datasets at any level. The next section will provide the results and performance of the developed system.

## III. Results and conclusion

The purpose of this study is to create a predictive algorithm to help hospitals and medical facilities maximize the number of survivors by providing an accurate and reliable tool to help medical decision making and triage COVID-19 patients more effectively and accurately during the pandemic.

As explained in section II, several metrics such as Accuracy, ROC, AUC, and Confusion Matrix have been used to evaluate the developed model.

Table 1 demonstrates the prediction accuracy for predicting mortality in patients with COVID-19 using 10-fold cross-validation for various machine learning algorithms.

**Table 1.**
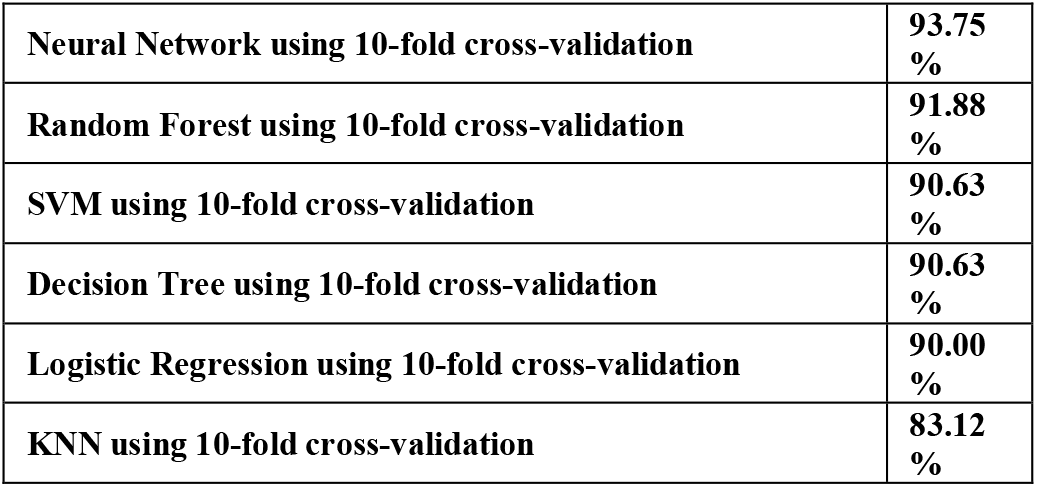
**The accuracy of mortality prediction in patients with COVID-19 using 10-fold cross-validation**.

|  |  |
| --- | --- |
| Neural Network using 10-fold cross-validation | 93.75 % |
| Random Forest using 10-fold cross-validation | 91.88 % |
| SVM using 10-fold cross-validation | 90.63 % |
| Decision Tree using 10-fold cross-validation | 90.63 % |
| Logistic Regression using 10-fold cross-validation | 90.00 % |
| KNN using 10-fold cross-validation | 83.12 % |

Figure 4 demonstrates and compares the ROC curves and AUC for every machine learning algorithm that was used in this research.

**Figure 4.**
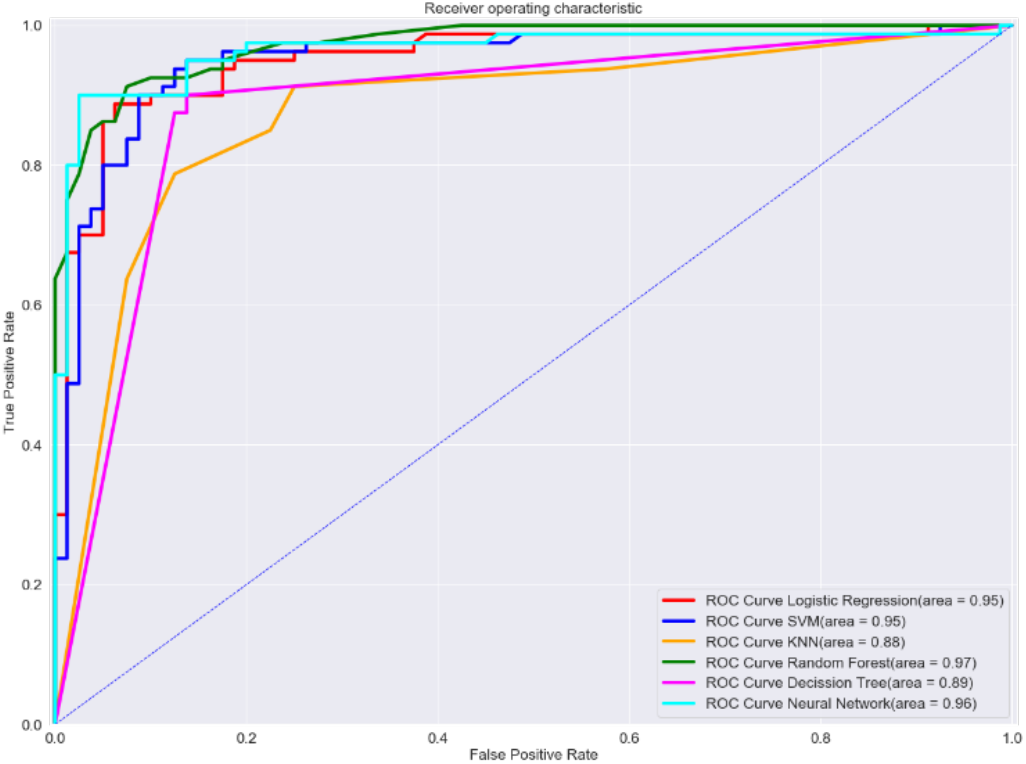
ROC Curve comparison for all algorithms

A confusion matrix (Figure 5) is used to describe and visualize the performance of the Neural Network algorithm classifier and also to provide insight on what the model misclassifies. The sensitivity and specificity of the model were calculated using the confusion matrix.

**Figure 5.**
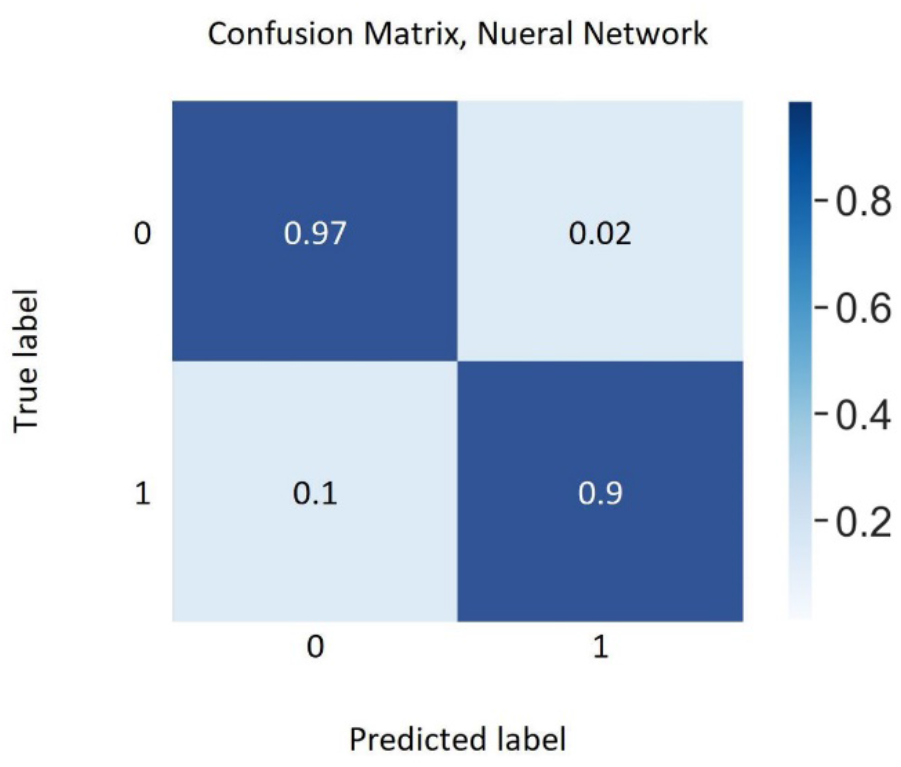
Neural Network confusion matrix

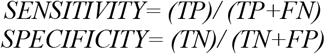

where TP: True positive, TN: True negative, FP: False positive, and FN: False negative.

The results demonstrate that the developed algorithm is able to accurately predict the mortality risk in patients with COVID-19 based on the patients’ physiological conditions, symptoms, and demographic information. Figure 6 shows the mortality risk predicted by the algorithm for sample patients.

**Figure 6.**
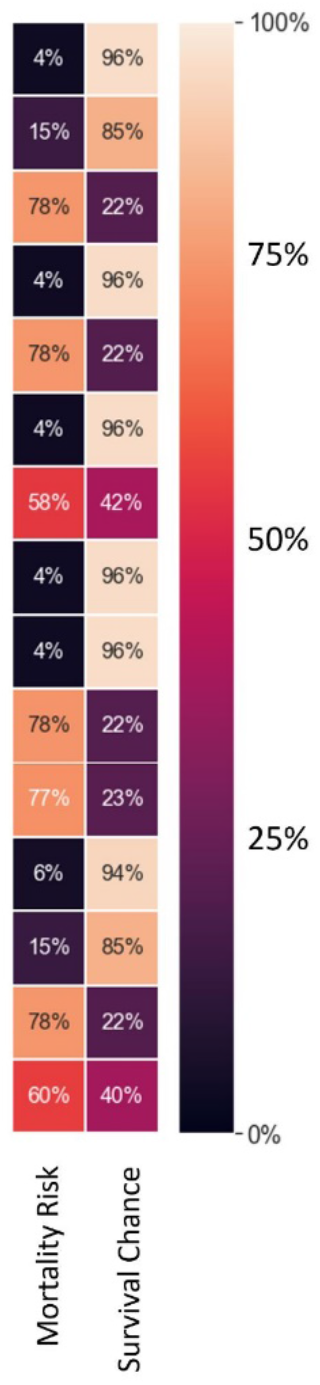
Mortality risk predicted for sample patients.

This system can help hospitals, medical facilities, and caregivers decide who needs to get attention first before other patients, triage patients when the system is overwhelmed by overcrowding, and also eliminate delays in providing the necessary care.

This study could expand to other diseases to help the healthcare system respond more effectively during an outbreak or a pandemic.

## Data Availability

The data used in this research is public.

